# Nicotine pouch, electronic cigarette and tobacco use and generalised anxiety among adolescents

**DOI:** 10.64898/2026.08.05.26359767

**Authors:** Otto Ruokolainen, Noora Berg, Jenni Helenius, Hanna Ollila, Olli Kiviruusu

## Abstract

**Background and Aims:** Anxiety remains prevalent among adolescents while tobacco and nicotine product use, especially the recent increases of novel product use such as e-cigarettes and nicotine pouches, raises further public health concerns. The associations between novel tobacco and nicotine product use and anxiety remains understudied. This study aims to determine whether tobacco and nicotine product use is associated with generalised anxiety and whether this association differs by used product.

**Design:** Cross-sectional survey, School Health Promotion study in 2025.

**Setting:** A school-based nationwide survey conducted in all Finnish lower and upper secondary schools.

**Participants:** Students aged 13–20 years in three school levels: 8.-9. grade students in lower secondary schools (N= 94 743, 73% of the students), 1st and 2nd year students in general upper secondary schools (n=47 248, 70% of the students) and of vocational institutions (n=24 998, 38% of the students).

**Measurements:** Exclusive (single product) and non-exclusive (≥1 products) daily or weekly use of tobacco and nicotine products, including nicotine pouches, e-cigarettes, cigarettes, and smokeless tobacco (snus). Generalised anxiety was measured using the Generalised Anxiety Disorder Scale (GAD-7). The cut-off of ≥10 points indicated participants with moderate to severe self-reported generalised anxiety symptoms. Background variables included sociodemographic variables and heavy drinking.

**Results:** Of the 166,989 participants 51.4% were females, mean age was 15.7 (SD 1.27), 21.2% reported generalised anxiety. Prevalence of generalised anxiety increased gradient-wise in accordance with both non-exclusive and exclusive use frequency of different tobacco and nicotine products, as well as with number of products used. Daily use of nicotine pouches was associated with higher odds of anxiety compared with never use (boys: adjusted odds ratios (aOR) 1.19, 95% CI, 1.05 to 1.34; girls: aOR 1.74, 95% CI, 1.58 to 1.91), yet the association between daily e-cigarette use seemed to be stronger (boys aOR 1.96, 95% CI, 1.54 to 2.49; girls: aOR 2.29, 95% CI, 2.09 to 2.51).

**Summary:** Any use of tobacco and nicotine products, including new products, is associated with generalised anxiety among adolescents, with some differences between products. Measures to prevent the initiation of tobacco and nicotine product use and to promote mental health among adolescents should be enacted.

## INTRODUCTION

Globally cigarette smoking prevalence has decreased among adolescents [1]. However, this positive trend is counteracted by a rapid increase in the availability and use of new tobacco and nicotine products (TNPs), such as e-cigarettes and nicotine pouches [2, 3]. While smoking has well-known health harms in addition to causing dependence, the literature of the health harms of new TNPs is also emerging [4–6].

Tobacco product use may worsen mental health via biological mechanisms (e.g. withdrawal symptoms such as anxiety and depression) [7]. In recent years, rates of depressive and anxiety symptoms among adolescents have risen, with the increase being particularly pronounced among females [8]. Adolescence is a sensitive period for brain development, which TNP use could infer by sensitising brains to, for example, other substance use [9]. Adolescents who have depressive and anxiety symptoms use TNPs more than those without symptoms. The association is well known regarding cigarette smoking [7, 10], and recent research confirms similar associations with electronic cigarette (e-cigarette) use (‘vaping’) [10–13]. Yet, the evidence is scarce for snus (Swedish type smokeless tobacco) [14, 15] and almost non-existent for oral nicotine pouches. In addition, multiple TNP use increases the rate of symptoms [10], indicating a possible dose-response association. Compared with girls, boys seem to use multiple TNPs (poly-use) more often [16].

There is a major gap in evidence considering the possible differences between exclusive (single product) and non-exclusive (≥1 products) use of TNPs and anxiety among adolescents, taking into account also new products such as nicotine pouches.

While the association between conventional cigarette smoking and mental health symptoms, including anxiety, is well established, there is a notable lack of research examining these associations across a broader spectrum of tobacco and nicotine products. In particular, evidence on newer products, such as nicotine pouches, remains extremely limited despite their rapidly increasing popularity among adolescents. Furthermore, little is known about the association between poly-use of tobacco and nicotine products and mental health symptoms. Examining these associations has important implications for preventing TNP use and mental health symptoms. The present study is, to our knowledge, the first one to analyse nicotine pouch use along with other TNPs – e-cigarettes, cigarettes, and snus – and the association of their use with mental health. Using large nationwide Finnish school survey in 2025, we addressed the following research questions: 1) How is the use of different tobacco and nicotine products, considering exclusive use of one product and non-exclusive use of at least one product, associated with generalised anxiety symptoms among adolescents? And 2) Do adolescents who use multiple tobacco or nicotine products (poly-use) report higher levels of generalised anxiety compared to those who use only one product or none at all?

## METHODS

### Data

The nationwide School Health Promotion (SHP) study is based on total sampling, i.e. all schools and all students in targeted grades in Finland are invited to participate in a survey study conducted biennially by the Finnish Institute for Health and Welfare (THL) [17]. This research is based on SHP 2025 that was collected between 3^rd^ March and 9^th^ May comprising 169,015 lower and upper secondary students. Unreliable responses (n=2026, 1.2%) based on implausible response patterns (for details see [17]) were excluded, leaving 166,989 responses for the analyses. Of these 94,743 were 8^th^ and 9^th^ grade students of lower secondary schools (73% coverage rate of all 8^th^ and 9^th^ grade students), 47,248 1^st^ and 2^nd^ year students in general upper secondary schools (70% coverage rate), and 24,998 1^st^ and 2^nd^ year students in vocational education institutions (38% coverage rate). The SHP survey is anonymous, confidential, and participation is voluntary. The participants completed the online questionnaire in a classroom setting, adolescents who were absent did not respond. Parents were informed about the study in advance and parents of children under the age of 15 had the right to prohibit the child’s participation. Responding to the survey was taken as informed consent to participate. The study protocol has been approved by the Institutional review board of THL. The procedures of the study were in accordance with the principles of the Helsinki Declaration on research with human subjects. A detailed description of the survey has been reported elsewhere [17] and details about the data collection on SHP Scientific Research Privacy notices [18]. This study adheres to the STROBE reporting guidelines.

### Measures

The questions and response options used in the measures are provided in Table S1. All the information is self-reported.

#### Exposure variables

TNP use was asked with a question: “How often have you used tobacco (cigarettes or cigars)/e-cigarettes/snus/nicotine pouches?” with the answer options “Never”, “I have tried a couple of times”, “I use less often than weekly”, “I use once a week or more often, but not daily”, “Daily”, and “I have quit using them”. Exclusive use of a TNP was created by removing those cases with at least weekly use of any other product from the original variable. A dichotomous variable for “weekly or more frequent use” was also created for each product type by combining categories “weekly but not daily use” and “daily use” (yes) vs. all other categories (no). Based on these variables a count of used TNPs was created (0, 1, 2, ≥3 products used at least weekly) and a variable comprising all possible combinations of different TNPs used at least weekly.

#### Dependent variable

Generalised anxiety was assessed with the Generalised Anxiety Disorder Scale (GAD-7) [19], asking how frequently respondents have experienced seven symptoms of generalised anxiety during the past two weeks, using four response options ranging from “Not at all” (0) to “Nearly every day” (3). When calculating the sum score, up to two missing items were allowed and replaced by the mean of the respondent’s other items. Participants scoring ≥10 points were defined as having moderate to severe generalised anxiety symptoms [19].

#### Background variables

The respondents reported their official gender (boy/girl), age (continuous, range 13–20 years), perception of their family’s financial situation (5-point scale from very good to very poor), living with both parents (yes/no), and family origin (“foreign background” if both parents were not born in Finland, otherwise “not foreign background”). School level consisted of lower secondary, general upper secondary, or vocational upper secondary school and heavy drinking was measured as monthly or more frequent consumption of alcohol until heavily drunk (yes/no).

Missing observations per variable varied between 0.0–6.3% (Table 1).

**Table 1.**
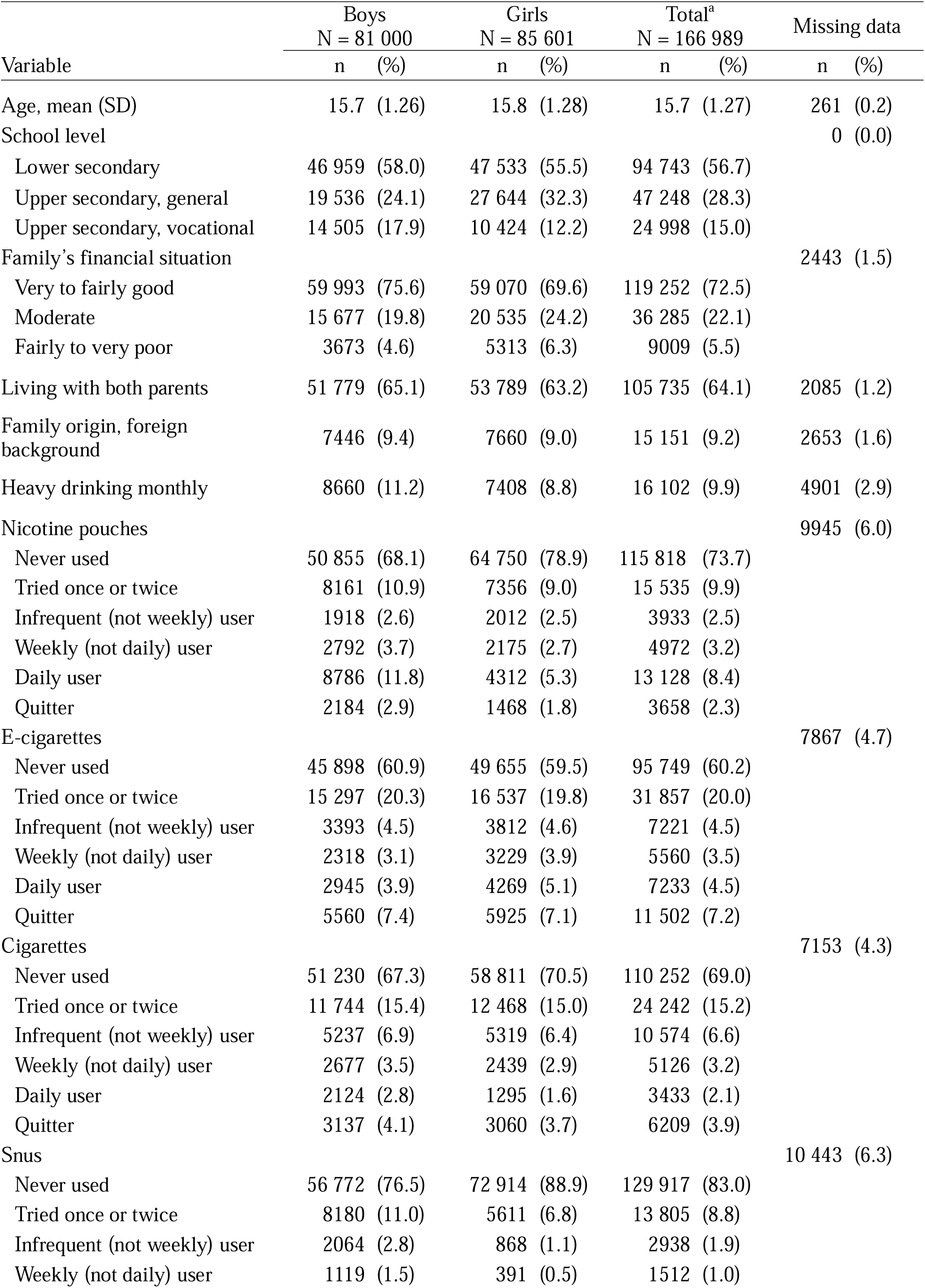
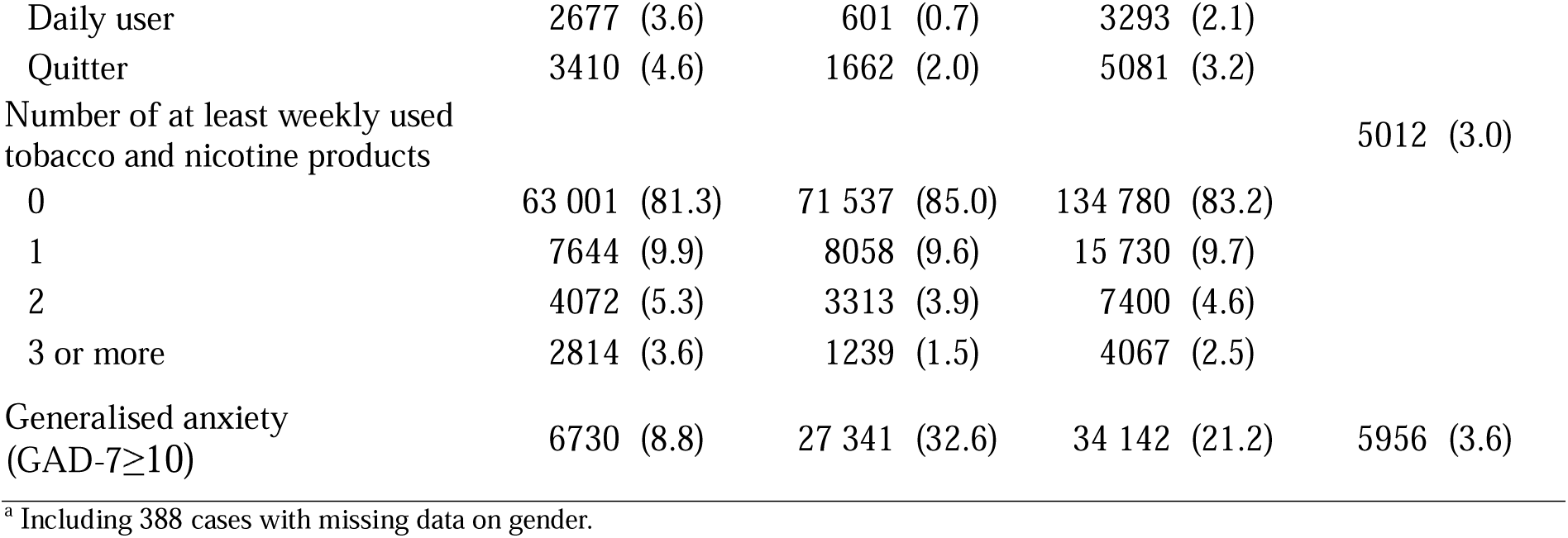
Distributions of the study variables by gender.

| Variable | Boys<br>N = 81 000 |  | Girls<br>N = 85 601 |  | Total<sup>a</sup><br>N = 166 989 |  | Missing data |  |
| --- | --- | --- | --- | --- | --- | --- | --- | --- |
|  | n | (%) | n | (%) | n | (%) | n | (%) |
| Age, mean (SD) | 15.7 | (1.26) | 15.8 | (1.28) | 15.7 | (1.27) | 261 | (0.2) |
| School level |  |  |  |  |  |  | 0 | (0.0) |
| Lower secondary | 46 959 | (58.0) | 47 533 | (55.5) | 94 743 | (56.7) |  |  |
| Upper secondary, general | 19 536 | (24.1) | 27 644 | (32.3) | 47 248 | (28.3) |  |  |
| Upper secondary, vocational | 14 505 | (17.9) | 10 424 | (12.2) | 24 998 | (15.0) |  |  |
| Family's financial situation |  |  |  |  |  |  | 2443 | (1.5) |
| Very to fairly good | 59 993 | (75.6) | 59 070 | (69.6) | 119 252 | (72.5) |  |  |
| Moderate | 15 677 | (19.8) | 20 535 | (24.2) | 36 285 | (22.1) |  |  |
| Fairly to very poor | 3673 | (4.6) | 5313 | (6.3) | 9009 | (5.5) |  |  |
| Living with both parents | 51 779 | (65.1) | 53 789 | (63.2) | 105 735 | (64.1) | 2085 | (1.2) |
| Family origin, foreign background | 7446 | (9.4) | 7660 | (9.0) | 15 151 | (9.2) | 2653 | (1.6) |
| Heavy drinking monthly | 8660 | (11.2) | 7408 | (8.8) | 16 102 | (9.9) | 4901 | (2.9) |
| Nicotine pouches |  |  |  |  |  |  | 9945 | (6.0) |
| Never used | 50 855 | (68.1) | 64 750 | (78.9) | 115 818 | (73.7) |  |  |
| Tried once or twice | 8161 | (10.9) | 7356 | (9.0) | 15 535 | (9.9) |  |  |
| Infrequent (not weekly) user | 1918 | (2.6) | 2012 | (2.5) | 3933 | (2.5) |  |  |
| Weekly (not daily) user | 2792 | (3.7) | 2175 | (2.7) | 4972 | (3.2) |  |  |
| Daily user | 8786 | (11.8) | 4312 | (5.3) | 13 128 | (8.4) |  |  |
| Quitter | 2184 | (2.9) | 1468 | (1.8) | 3658 | (2.3) |  |  |
| E-cigarettes |  |  |  |  |  |  | 7867 | (4.7) |
| Never used | 45 898 | (60.9) | 49 655 | (59.5) | 95 749 | (60.2) |  |  |
| Tried once or twice | 15 297 | (20.3) | 16 537 | (19.8) | 31 857 | (20.0) |  |  |
| Infrequent (not weekly) user | 3393 | (4.5) | 3812 | (4.6) | 7221 | (4.5) |  |  |
| Weekly (not daily) user | 2318 | (3.1) | 3229 | (3.9) | 5560 | (3.5) |  |  |
| Daily user | 2945 | (3.9) | 4269 | (5.1) | 7233 | (4.5) |  |  |
| Quitter | 5560 | (7.4) | 5925 | (7.1) | 11 502 | (7.2) |  |  |
| Cigarettes |  |  |  |  |  |  | 7153 | (4.3) |
| Never used | 51 230 | (67.3) | 58 811 | (70.5) | 110 252 | (69.0) |  |  |
| Tried once or twice | 11 744 | (15.4) | 12 468 | (15.0) | 24 242 | (15.2) |  |  |
| Infrequent (not weekly) user | 5237 | (6.9) | 5319 | (6.4) | 10 574 | (6.6) |  |  |
| Weekly (not daily) user | 2677 | (3.5) | 2439 | (2.9) | 5126 | (3.2) |  |  |
| Daily user | 2124 | (2.8) | 1295 | (1.6) | 3433 | (2.1) |  |  |
| Quitter | 3137 | (4.1) | 3060 | (3.7) | 6209 | (3.9) |  |  |
| Snus |  |  |  |  |  |  | 10 443 | (6.3) |
| Never used | 56 772 | (76.5) | 72 914 | (88.9) | 129 917 | (83.0) |  |  |
| Tried once or twice | 8180 | (11.0) | 5611 | (6.8) | 13 805 | (8.8) |  |  |
| Infrequent (not weekly) user | 2064 | (2.8) | 868 | (1.1) | 2938 | (1.9) |  |  |
| Weekly (not daily) user | 1119 | (1.5) | 391 | (0.5) | 1512 | (1.0) |  |  |

|  |  |  |  |  |
| --- | --- | --- | --- | --- |
| Daily user | 2677 (3.6) | 601 (0.7) | 3293 (2.1) |  |
| Quitter | 3410 (4.6) | 1662 (2.0) | 5081 (3.2) |  |
| Number of at least weekly used tobacco and nicotine products |  |  |  | 5012 (3.0) |
| 0 | 63 001 (81.3) | 71 537 (85.0) | 134 780 (83.2) |  |
| 1 | 7644 (9.9) | 8058 (9.6) | 15 730 (9.7) |  |
| 2 | 4072 (5.3) | 3313 (3.9) | 7400 (4.6) |  |
| 3 or more | 2814 (3.6) | 1239 (1.5) | 4067 (2.5) |  |
| Generalised anxiety (GAD-7≥10) | 6730 (8.8) | 27 341 (32.6) | 34 142 (21.2) | 5956 (3.6) |

<sup>a</sup> Including 388 cases with missing data on gender.

### Statistical analyses

All analyses were conducted using SPSS version 31.0 and performed separately for boys and girls due to known gender differences in the prevalence of anxiety and TNP use. Missing data were handled using listwise deletion, applied separately for each analysis.

First, the frequencies of TNP use, as well as the number and combinations of at least weekly TNP use, were calculated. The use of TNPs and generalised anxiety were cross tabulated to analyse their associations, using both the original TNP variables (including use of a TNP irrespective of other TNP use) and exclusive use of a TNP (no other TNP use). Online-only supplements presents these results stratified by school level (lower and upper secondary).

Associations between exclusive use of TNPs and anxiety were then analysed with logistic regression models adjusting for all background factors. Associations between the number of TNPs and anxiety were also analysed using logistic regression models, presenting both unadjusted and adjusted (for all background factors) estimates with 95% confidence intervals. Two sets of models were run with both zero TNPs and one TNP as the reference category. Finally, logistic regression models were run on the association between combinations of at least weekly used TNPs and anxiety. No pre-registration was done so the findings should be described as exploratory.

## RESULTS

Girls represented 51.4% of the sample, mean age of respondents was 15.7 years (SD 1.27), and 21.2% reported generalised anxiety (Table 1). Most respondents had never used TNPs, ranging from 60.2% (e-cigarettes) to 83.0% (snus). The most commonly used products on a daily or weekly basis were nicotine pouches among boys (15.5%) and e-cigarettes among girls (9.0%). About 2% of participants smoked daily. One in ten respondents used only one TNP, while 7.1% used two or more TNPs. The most commonly reported product combination was “nicotine pouches only” (6.9%) among boys, and “e-cigarettes only” (4.7%) among girls, while 1.6% of boys and 0.4% of girls used all four TNPs (Table S2). Among both boys and girls, the most common poly-use combination was the use of e-cigarettes and nicotine pouches (boys: 1.9%, girls: 1.8%).

The prevalence of generalised anxiety increased in a gradient-like manner with both the frequency of exclusive and non-exclusive use of different TNPs (Table 2). Overall, anxiety was most prevalent among adolescents who used TNPs daily and least prevalent among those who had never used TNPs. About half of the girls who used TNPs daily reported generalised anxiety, whereas the prevalence among boys was markedly lower, typically below 20%. Overall, daily smoking and e-cigarette use seemed to have somewhat stronger associations with anxiety than daily nicotine pouch or snus use, especially among boys.

**Table 2.** Percentages of those with generalised anxiety (GAD-7≥10) by use of different tobacco and nicotine products.

| Subgroup/tobacco and nicotine product |  |  | Never used |  | Tried once or twice |  | Infrequent (not weekly) user |  | Weekly (not daily) user |  | Daily user |  | Quitter |  |
| --- | --- | --- | --- | --- | --- | --- | --- | --- | --- | --- | --- | --- | --- | --- |
|  |  |  | n | (%) | n | (%) | n | (%) | n | (%) | n | (%) | n | (%) |
| **Boys** |  |  |  |  |  |  |  |  |  |  |  |  |  |  |
| Nicotine pouches | all<sup>b</sup> | 23 841 | 3817 | (7.6) | 642 | (8.0) | 216 | (11.5) | 319 | (11.6) | 1095 | (12.9) | 256 | (12.0) |
|  | exclusive<sup>c</sup> | 15 764 | 3720 | (7.5) | 524 | (7.2) | 103 | (8.7) | 100 | (8.1) | 372 | (9.4) | 197 | (11.1) |
| E-cigarettes | all<sup>b</sup> | 29 513 | 3285 | (7.3) | 1225 | (8.1) | 367 | (11.1) | 317 | (14.1) | 598 | (21.5) | 685 | (12.6) |
|  | exclusive<sup>c</sup> | 17 586 | 3182 | (7.2) | 1011 | (8.0) | 139 | (11.4) | 80 | (15.9) | 97 | (17.9) | 328 | (13.5) |
| Cigarettes | all<sup>b</sup> | 24 919 | 3711 | (7.3) | 967 | (8.4) | 578 | (11.3) | 360 | (13.9) | 427 | (21.6) | 426 | (13.9) |
|  | exclusive<sup>c</sup> | 13 026 | 3584 | (7.2) | 718 | (8.2) | 166 | (9.9) | 67 | (11.9) | 46 | (16.3) | 215 | (15.1) |
| Snus | all<sup>b</sup> | 17 450 | 4334 | (7.7) | 752 | (9.3) | 233 | (11.5) | 172 | (15.9) | 429 | (16.8) | 411 | (12.3) |
|  | exclusive<sup>c</sup> | 6993 | 3984 | (7.5) | 407 | (8.1) | 39 | (10.4) | 19 | (21.1) | 33 | (13.5) | 131 | (11.8) |
| **Girls** |  |  |  |  |  |  |  |  |  |  |  |  |  |  |
| Nicotine pouches | all<sup>b</sup> | 17 323 | 18 987 | (29.6) | 2840 | (38.8) | 889 | (44.7) | 1012 | (47.0) | 2128 | (49.9) | 683 | (47.2) |
|  | exclusive<sup>c</sup> | 10 078 | 18 027 | (29.0) | 1866 | (34.9) | 288 | (36.8) | 299 | (37.7) | 929 | (43.9) | 408 | (41.8) |
| E-cigarettes | all<sup>b</sup> | 33 772 | 13 463 | (27.3) | 5639 | (34.3) | 1541 | (40.7) | 1524 | (47.7) | 2305 | (54.9) | 2529 | (43.0) |
|  | exclusive<sup>c</sup> | 25 446 | 13 369 | (27.3) | 5094 | (33.6) | 904 | (37.5) | 748 | (45.0) | 1131 | (50.9) | 1600 | (42.1) |
| Cigarettes | all<sup>b</sup> | 24 581 | 16 624 | (28.5) | 4539 | (36.7) | 2355 | (44.6) | 1227 | (51.0) | 675 | (53.4) | 1467 | (48.4) |
|  | exclusive<sup>c</sup> | 14 429 | 16 127 | (28.2) | 3263 | (34.2) | 829 | (37.9) | 298 | (43.3) | 173 | (46.6) | 718 | (46.6) |
| Snus | all<sup>b</sup> | 9133 | 22 432 | (31.0) | 2328 | (41.8) | 433 | (50.4) | 204 | (53.5) | 298 | (50.9) | 796 | (48.4) |
|  | exclusive<sup>c</sup> | 3409 | 19 471 | (29.4) | 938 | (34.5) | 53 | (43.4) | 14 | (38.9) | 29 | (36.3) | 179 | (42.4) |

<sup>a</sup> Number of cases ever used the given tobacco and nicotine product (including those tried once or twice, using currently, and quitters).
<sup>b</sup> Including all cases using the product.
<sup>c</sup> Excluding those using weekly or daily any other tobacco and nicotine products.

Table 3 presents adjusted odds ratios for the association between exclusive TNP use and generalised anxiety. Compared with never use, having tried TNPs and their infrequent use was predominantly associated with higher adjusted odds of generalised anxiety, and the association seemed stronger for girls. The associations mostly grew stronger when moving to more frequent use. Daily use of all the different products, except snus, were associated with generalised anxiety among boys and girls.

**Table 3.** Adjusted^a^ logistic regression models predicting generalised anxiety (GAD-7≥10) by exclusive use of different tobacco and nicotine products.

| Subgroup/tobacco and nicotine product | Never used (ref.) | Tried once or twice |  | Infrequent (not weekly) user |  | Weekly (not daily) user |  | Daily user |  | Quitter |  |
| --- | --- | --- | --- | --- | --- | --- | --- | --- | --- | --- | --- |
|  | OR | OR | (95% CI) | OR | (95% CI) | OR | (95% CI) | OR | (95% CI) | OR | (95% CI) |
| **Boys** |  |  |  |  |  |  |  |  |  |  |  |
| Nicotine pouches<sup>b</sup> | 1.00 | 0.96 | (0.87–1.06) | 1.16 | (0.94–1.43) | 1.05 | (0.84–1.30) | 1.19 | (1.05–1.34) | 1.45 | (1.24–1.71) |
| E-cigarettes<sup>b</sup> | 1.00 | 1.10 | (1.02–1.18) | 1.45 | (1.20–1.75) | 1.98 | (1.52–2.57) | 1.96 | (1.54–2.49) | 1.84 | (1.62–2.10) |
| Cigarettes<sup>b</sup> | 1.00 | 1.08 | (0.99–1.18) | 1.25 | (1.05–1.48) | 1.27 | (0.95–1.68) | 1.75 | (1.24–2.47) | 1.93 | (1.65–2.27) |
| Snus<sup>b</sup> | 1.00 | 1.04 | (0.93–1.16) | 1.26 | (0.89–1.78) | 2.37 | (1.35–4.14) | 1.48 | (0.99–2.22) | 1.46 | (1.20–1.77) |
| **Girls** |  |  |  |  |  |  |  |  |  |  |  |
| Nicotine pouches<sup>b</sup> | 1.00 | 1.27 | (1.19–1.35) | 1.38 | (1.19–1.61) | 1.42 | (1.22–1.65) | 1.74 | (1.58–1.91) | 1.60 | (1.40–1.82) |
| E-cigarettes<sup>b</sup> | 1.00 | 1.32 | (1.26–1.37) | 1.50 | (1.38–1.64) | 1.94 | (1.75–2.15) | 2.29 | (2.09–2.51) | 1.80 | (1.68–1.93) |
| Cigarettes<sup>b</sup> | 1.00 | 1.28 | (1.22–1.35) | 1.46 | (1.33–1.60) | 1.72 | (1.47–2.01) | 1.89 | (1.53–2.34) | 1.99 | (1.79–2.22) |
| Snus<sup>b</sup> | 1.00 | 1.17 | (1.07–1.27) | 1.62 | (1.12–2.35) | 0.96 | (0.46–1.99) | 1.09 | (0.68–1.74) | 1.55 | (1.27–1.89) |

<sup>a</sup> Adjusted for age, school level, family's financial situation, living with both parents, foreign background origin, heavy drinking monthly.
<sup>b</sup> Excluding those using weekly or daily other tobacco and nicotine products.

While daily use of nicotine pouches was associated with increased odds of anxiety (boys: aOR 1.19, 95% CI 1.05–1.34; girls: aOR 1.74, 95% CI 1.58–1.91), the association between daily vaping appeared stronger among both genders (boys: aOR 1.96, 95% CI 1.54–2.49; girls: aOR 2.29, 95% CI 2.09–2.51).

Out of different product combinations of use, the association with anxiety was most pronounced for “e-cigarettes + cigarettes + snus” use among boys and “nicotine pouches + e-cigarettes + snus” use among girls (Table 4). The most prevalent poly-use combination, i.e. “nicotine pouches + e-cigarettes” had an adjusted odds ratio of 1.73 (95% CI 1.48–2.02) for anxiety among boys and 2.33 (95% CI 2.09–2.59) among girls.

**Table 4.** Generalised anxiety (GAD-7≥10) by combinations of different tobacco and nicotine products used at least weekly.

| Combination of tobacco and nicotine products | Boys |  |  |  | Girls |  |  |  |
| --- | --- | --- | --- | --- | --- | --- | --- | --- |
|  | n | (%) | OR<sup>a</sup> | (95% CI) | n | (%) | OR<sup>a</sup> | (95% CI) |
| No weekly or daily use | 4765 | (7.7) | 1.00 |  | 21 116 | (29.8) | 1.00 |  |
| Nicotine pouches only | 472 | (9.0) | 1.14 | (1.03–1.27) | 1228 | (42.2) | 1.60 | (1.48–1.73) |
| E-cigarettes only | 177 | (17.0) | 1.86 | (1.56–2.22) | 1879 | (48.4) | 1.87 | (1.75–2.00) |
| Cigarettes only | 113 | (13.4) | 1.42 | (1.14–1.76) | 471 | (44.4) | 1.63 | (1.43–1.85) |
| Snus only | 52 | (15.6) | 1.81 | (1.31–2.50) | 43 | (37.1) | 1.09 | (0.74–1.62) |
| Nicotine pouches + e-cigarettes | 217 | (15.0) | 1.73 | (1.48–2.02) | 827 | (54.9) | 2.33 | (2.09–2.59) |
| Nicotine pouches + cigarettes | 104 | (12.4) | 1.42 | (1.14–1.77) | 290 | (50.8) | 1.91 | (1.61–2.26) |
| Nicotine pouches + snus | 116 | (10.2) | 1.27 | (1.04–1.56) | 107 | (44.6) | 1.66 | (1.27–2.16) |
| E-cigarettes + cigarettes | 76 | (20.2) | 2.15 | (1.64–2.81) | 489 | (54.8) | 2.24 | (1.95–2.60) |
| E-cigarettes + snus | 13 | (18.3) | 1.09 | (0.54–2.20) | 21 | (53.8) | 2.30 | (1.20–4.40) |
| Cigarettes + snus | 24 | (22.0) | 2.02 | (1.21–3.36) | 14 | (46.7) | 1.62 | (0.77–3.38) |
| Nicotine pouches + e-cigarettes + cigarettes | 121 | (17.8) | 1.97 | (1.59–2.43) | 378 | (56.8) | 2.14 | (1.82–2.52) |
| Nicotine pouches + e-cigarettes + snus | 47 | (17.5) | 1.88 | (1.34–2.63) | 57 | (62.6) | 3.14 | (2.03–4.84) |
| Nicotine pouches + cigarettes + snus | 85 | (15.0) | 1.68 | (1.31–2.16) | 82 | (59.4) | 2.56 | (1.80–3.64) |
| E-cigarettes + cigarettes + snus | 12 | (33.3) | 4.04 | (1.90–8.59) | 7 | (50.0) | 2.06 | (0.71–5.95) |
| Nicotine pouches + e-cigarettes + cigarettes + snus | 252 | (22.7) | 2.24 | (1.90–2.64) | 171 | (57.2) | 2.23 | (1.75–2.84) |

<sup>a</sup> Models adjusted for age, school level, family's financial situation, living with both parents, foreign background origin, heavy drinking monthly.

In addition to the frequency of use, the associations between number of used TNPs and anxiety seemed gradient-like (Table 5). Use of at least one TNP was associated with elevated odds of anxiety compared with not using TNPs (aORs ranging from 1.31 to 2.03 among boys, from 1.73 to 2.28 among girls). Furthermore, using at least two products was associated with higher odds of anxiety compared with using just one product, in unadjusted and adjusted models.

**Table 5.** Association between the number of tobacco and nicotine products used at least weekly and generalised anxiety (GAD-7≥10).

| Subgroup/number of products | n | (%) | Models with 0 products as the reference |  | Models with 1 product as the reference |  |
| --- | --- | --- | --- | --- | --- | --- |
|  |  |  | OR<sup>a</sup> (95% CI) | OR<sup>b</sup> (95% CI) | OR<sup>a</sup> (95% CI) | OR<sup>b</sup> (95% CI) |
| **Boys** |  |  |  |  |  |  |
| 0 products | 4765 | (7.7) | 1.00 | 1.00 | 0.68 (0.63–0.73) | 0.77 (0.70–0.83) |
| 1 product | 814 | (10.9) | 1.48 (1.37–1.60) | 1.31 (1.20–1.43) | 1.00 | 1.00 |
| 2 products | 550 | (13.9) | 1.93 (1.76–2.12) | 1.57 (1.42–1.75) | 1.31 (1.17–1.47) | 1.20 (1.07–1.36) |
| 3 or more products | 517 | (19.4) | 2.90 (2.62–3.20) | 2.03 (1.81–2.28) | 1.96 (1.74–2.21) | 1.55 (1.36–1.77) |
| **Girls** |  |  |  |  |  |  |
| 0 products | 21 116 | (29.8) | 1.00 | 1.00 | 0.51 (0.49–0.53) | 0.58 (0.55–0.61) |
| 1 product | 3621 | (45.4) | 1.96 (1.87–2.06) | 1.73 (1.64–1.82) | 1.00 | 1.00 |
| 2 products | 1748 | (53.3) | 2.69 (2.51–2.89) | 2.17 (2.01–2.34) | 1.37 (1.27–1.49) | 1.26 (1.16–1.37) |
| 3 or more products | 695 | (57.5) | 3.20 (2.85–3.59) | 2.28 (2.01–2.57) | 1.63 (1.44–1.84) | 1.32 (1.16–1.50) |

<sup>a</sup> Unadjusted models.
<sup>b</sup> Models adjusted for age, school level, family's financial situation, living with both parents, foreign background origin, heavy drinking monthly.

Stratified analyses of different school levels on the association between TNP use and anxiety showed that prevalence of anxiety was somewhat on a similar level among younger and older adolescents who had never used TNPs while anxiety among adolescents who used TNPs daily was generally more common among younger adolescents (Table S3).

## DISCUSSION

Our results of a large nationwide study of adolescents aged 13–20 years showed that the use of each of the studied tobacco and nicotine product (TNP) was associated with generalised anxiety. Overall, the associations showed a gradient-like pattern in relation to the frequency of use and the number of products used. There was variation in the associations between different TNP combinations and anxiety, and the association between TNP use and anxiety seemed stronger for girls in some cases.

Our results support previous strong evidence of a positive association between smoking and mental health symptoms and more limited, yet aligned, evidence of a positive association between e-cigarette use and mental health symptoms [7, 10, 20]. Furthermore, our study provides novel insights into the association between nicotine pouch use and mental health symptoms in a context where such use has increased substantially among Finnish adolescents and is more prevalent among them than among their peers in countries such as the USA and Great Britain [21–24]. Daily nicotine pouch use was associated with generalised anxiety among boys and girls, while weekly nicotine pouch use was associated with anxiety only among girls. Among girls, the association between daily use of nicotine pouches seemed to be somewhat as strong as with daily smoking and anxiety, while daily e-cigarette use showed a stronger association with anxiety. Notably, even less frequent nicotine pouch use was associated with elevated odds of anxiety among girls. These findings propose that the association between TNP use and anxiety is stronger among girls than among boys. This might be due to more frequent use of nicotine pouches among boys (thus more clearly distinguishing girls with high-risk use) or to gender differences in the comorbidity patterns between nicotine pouch use and anxiety. In addition, females are generally more likely to report internalising mental health symptoms, which has been attributed to gendered responses to negative affect [25]. Future studies should investigate the pathways of different TNP use and anxiety in more detail with longitudinal study designs.

The current study found inconclusive association between daily snus use and anxiety, both supporting and contradicting previous findings from Nordic countries. A Swedish study on 17-year-old adolescents found inconclusive association between exclusive snus use and depressive symptoms [15] while a prior study on Norwegian college and university students found a positive association between snus use and mental health symptoms [14]. Differences in findings may be explained by variations in study populations and mental health measures, as, for example, the Norwegian study included older participants (18–25 years), used a broader symptoms measure encompassing both anxiety and depressive symptoms, and had higher prevalence of snus use alongside a rather low online survey response rate.

In addition to the finding that using one TNP is associated with higher odds of anxiety compared with not using TNPs, the current study observed that using two or more TNPs was associated with higher odds of anxiety compared with using only one product. These results imply a dose-response association with TNP use and mental health symptoms [26, 27]. Our results are in accordance with previous results showing that poly-use of TNPs is more common than exclusive use of TNPs among adolescents [28]. This highlights the challenges in the current tobacco and nicotine market, where the number of different products has increased^2^, in relation to adolescent mental health.

Different TNP combinations had varying association with anxiety and these were different for boys and girls; the strongest association was observed of poly-use of three products where smoking was included among boys (smoking, snus use and vaping) but was absent among girls (vaping, snus use, nicotine pouch use). This further highlights the differences between TNP use and mental health symptoms between genders. The use of new TNPs, such as nicotine pouches or snus, may increase the nicotine levels in the body more than cigarette use [29, 30]. It has been indicated that it is rather common for people who use the products not to know the nicotine strength of the used snus or nicotine pouches [31]. Limiting nicotine strengths in TNPs would be advisable also to prevent the harms of use among adolescents, with and without mental health symptoms.

Our results propose that anxiety is more pronounced among adolescents who used TNPs in lower secondary school than in upper secondary education. Nicotine pouch use was clearly associated with anxiety among boys in lower secondary education, but not among those in upper secondary education. Future studies should examine whether differences in the observed association are explained by age, school environment or some other underlying factors. However, based on the current results, younger adolescents seem to be more vulnerable when looking at the interplay between TNP use and anxiety.

The tobacco industry plays a decisive role by continuing to promote TNPs to adolescents. Adolescents are exposed to TNPs especially in different digital media platforms even in jurisdictions where strong tobacco control measures prevent exposure in traditional media environments [32]. As adolescents with mental health symptoms use social media more than other adolescents [33], they could be exposed to different TNPs more, possibly leading to widening differences on the use of these products. Thus, future measures to prevent adolescent TNP use should include more rigorous regulation of the digital environment.

The association between TNP use and mental health may be bi-directional, yet evidence is scarce and the role of tobacco and nicotine constituents remains contested [34, 35]. Our study examined comorbidity between TNP use and mental health symptoms while longitudinal studies are needed to examine the possible causal pathways. Our results emphasise the need to monitor different TNP use, including its combinations, and possible changes in it among adolescents, especially among those with mental health symptoms. Early identification, and support for tobacco and nicotine use cessation, in pediatric and student health services may provide opportunities to address the comorbidities before the onset of disorders.

### Limitations and strengths

The study relies on self-reported data which might introduce bias. However, the anonymous survey design may reduce misreporting, and unreliable responses were excluded. The results may not fully represent all students, particularly those who were absent from school. Furthermore, the coverage rate was lower in vocational schools than in lower secondary and general upper secondary schools, which warrants caution when generalising the findings to the target population as a whole.

Nevertheless, it is plausible that higher coverage would have resulted in even higher observed rates of TNP use. Finally, the cross-sectional design inhibits any causal interpretations.

Strengths include a large, nationwide sample of more than 160,000 respondents. We assessed both tobacco and new nicotine product use using a multiclass measure, providing a more detailed information than, for example, a dichotomous approach. GAD-7 [19] is validated and internationally widely used measure for generalised anxiety. Furthermore, to our knowledge this is the first large-scale study to examine the association between nicotine pouch use and anxiety among adolescents.

## CONCLUSIONS

The study shows a clear association with the use of different tobacco and nicotine products, including new products like e-cigarettes and nicotine pouches, and generalised anxiety. Poly-use of different tobacco and nicotine products was more strongly associated with anxiety than using only one product. Nicotine pouch use showed a stronger association with anxiety among girls than among boys, while e-cigarette use was more strongly associated with anxiety than nicotine pouch use. Future measures to prevent tobacco and nicotine product use and promote mental health among adolescents are needed.

## Data Availability

Finnish Institute for Health and Welfare (THL) has collected the data and has the rights for the data. THL is committed to complying with the established code of conduct in its field. THL produces public statistical reports and interactive reports, but the data is confidential. Researchers can apply the data from Finnish Social Science Data Archive (https://www.fsd.tuni.fi/en/).

