## Supplementary Material for "Nicotine pouch, electronic cigarette and tobacco use and generalised anxiety among adolescents"

1. Table S1. Questions and response options for the measures of the study
2. Table S2. Combinations of different tobacco and nicotine products used at least weekly
3. Table S3. Percentages of those with generalised anxiety (GAD-7≥10) by use of different tobacco and nicotine products among lower and upper secondary students.

**Table S1. Questions and response options for the measures of the study.**

| Measure | Question | Response options |
|---|---|---|
| Tobacco and nicotine product use | How often have you used tobacco (cigarettes or cigars)/e-cigarettes/snus/nicotine pouches? | Never<br>I have tried a couple of times<br>I use less often than weekly<br>I use once a week or more often, but not daily<br>Daily<br>I have quit using them |
| Generalised Anxiety Disorder Scale (GAD-7) | Over the last two weeks, how often have you been bothered by any of the following problems?<br>The items: 'Feeling nervous, anxious or on edge, 'Not being able to stop or control worrying', 'Worrying too much about different things', 'Having trouble relaxing', 'Being so restless that it is hard to sit still', 'Becoming easily annoyed or irritable', 'Feeling afraid, as if something awful might happen'. | Not at all<br>Several days<br>More than half the days<br>Nearly every day |
| Gender | What is your official gender? | Boy<br>Girl |
| Age | How old are you now? I have turned | Answer options (ages) depending on the school level and type:<br>13 years or I am younger<br>14 years<br>15 years<br>16 years<br>17 years<br>18 years<br>19 years<br>20 years or I am older |
| Family's financial situation | What do you think your family's financial situation is like? | Very good<br>Fairly good<br>Moderate<br>Fairly poor<br>Very poor |
| Living arrangements | Where do you live? Select the option that best describes your situation. | In a shared home with my parents<br>Alternately with my parents<br>With one of my parents<br>In a foster family<br>At a children's home, a youth home or a reform school<br>In a professional foster home<br>Elsewhere |
| Family foreign background origin | In which country were you and your parents born? Please answer for everyone.<br><br>You<br>Mother<br>Father | Finland<br>Sweden<br>Estonia<br>Russia<br>Ukraine<br>Other European country<br>Somalia<br>Syria<br>Iraq |

| | | |
|---|---|---|
| | | Iran<br>Afghanistan<br>India<br>China<br>Thailand<br>Vietnam<br>Some other country |
| Heavy drinking | How often do you drink alcohol so that you are heavily drunk? | Once a week or more often<br>About 1 to 2 times a month<br>Less frequently<br>Never |

**Table S2. Combinations of different tobacco and nicotine products used at least weekly.**

| | Boys | | Girls | |
|---|---|---|---|---|
| Combination of tobacco and nicotine products | n | (%) | n | (%) |
| No weekly or daily use | 63 001 | (81.3) | 71 537 | (85.0) |
| Nicotine pouches only | 5314 | (6.9) | 2923 | (3.5) |
| E-cigarettes only | 1086 | (1.4) | 3932 | (4.7) |
| Cigarettes only | 899 | (1.2) | 1082 | (1.3) |
| Snus only | 345 | (0.4) | 121 | (0.1) |
| Nicotine pouches + e-cigarettes | 1479 | (1.9) | 1524 | (1.8) |
| Nicotine pouches + cigarettes | 858 | (1.1) | 574 | (0.7) |
| Nicotine pouches + snus | 1153 | (1.5) | 241 | (0.3) |
| E-cigarettes + cigarettes | 392 | (0.5) | 905 | (1.1) |
| E-cigarettes + snus | 77 | (0.1) | 39 | (0.0) |
| Cigarettes + snus | 113 | (0.1) | 30 | (0.0) |
| Nicotine pouches + e-cigarettes + cigarettes | 706 | (0.9) | 678 | (0.8) |
| Nicotine pouches + e-cigarettes + snus | 275 | (0.4) | 96 | (0.1) |
| Nicotine pouches + cigarettes + snus | 585 | (0.8) | 141 | (0.2) |
| E-cigarettes + cigarettes + snus | 40 | (0.1) | 14 | (0.0) |
| Nicotine pouches + e-cigarettes + cigarettes + snus | 1208 | (1.6) | 310 | (0.4) |

**Table S3. Percentages of those with generalised anxiety (GAD-7≥10) by use of different tobacco and nicotine products among lower and upper secondary students.**

| Subgroup/tobacco and nicotine product | | n of users[a] | Never used | | Tried once or twice | | Infrequent (not weekly) user | | Weekly (not daily) user | | Daily user | | Quitter | |
|---|---|---|---|---|---|---|---|---|---|---|---|---|---|---|
| | | | n | (%) | n | (%) | n | (%) | n | (%) | n | (%) | n | (%) |
| **Lower secondary** | | | | | | | | | | | | | | |
| Boys | | | | | | | | | | | | | | |
| Nicotine pouches | all[b] | 11 733 | 2320 | (7.6) | 376 | (9.1) | 139 | (15.0) | 208 | (15.4) | 647 | (17.3) | 171 | (14.5) |
| | exclusive[c] | 7415 | 2276 | (7.5) | 308 | (8.1) | 64 | (11.5) | 60 | (11.0) | 177 | (13.0) | 126 | (13.0) |
| E-cigarettes | all[b] | 15 269 | 1993 | (7.2) | 692 | (9.3) | 207 | (14.4) | 210 | (16.4) | 437 | (23.4) | 418 | (15.1) |
| | exclusive[c] | 9680 | 1947 | (7.1) | 605 | (9.0) | 88 | (14.5) | 55 | (17.5) | 64 | (19.9) | 217 | (14.5) |
| Cigarettes | all[b] | 11 302 | 2375 | (7.4) | 514 | (10.0) | 324 | (16.1) | 198 | (16.9) | 274 | (25.6) | 257 | (17.4) |
| | exclusive[c] | 5771 | 2298 | (7.3) | 357 | (9.1) | 86 | (14.5) | 32 | (13.5) | 20 | (22.0) | 125 | (16.5) |
| Snus | all[b] | 8016 | 2666 | (7.9) | 419 | (11.3) | 138 | (15.9) | 99 | (19.9) | 282 | (23.2) | 244 | (16.8) |
| | exclusive[c] | 3268 | 2450 | (7.5) | 220 | (9.4) | 27 | (16.6) | 13 | (29.5) | 14 | (18.7) | 90 | (16.3) |
| Girls | | | | | | | | | | | | | | |
| Nicotine pouches | all[b] | 7685 | 11 497 | (30.8) | 1446 | (45.0) | 509 | (50.9) | 575 | (54.9) | 938 | (58.4) | 392 | (53.0) |
| | exclusive[c] | 3834 | 10 977 | (30.2) | 921 | (40.9) | 140 | (46.8) | 110 | (43.1) | 279 | (53.4) | 230 | (47.3) |
| E-cigarettes | all[b] | 16 055 | 8424 | (28.2) | 2840 | (38.3) | 705 | (45.7) | 896 | (51.8) | 1538 | (59.4) | 1255 | (48.2) |
| | exclusive[c] | 12 602 | 8394 | (28.2) | 2705 | (37.9) | 485 | (44.2) | 473 | (50.2) | 725 | (55.4) | 926 | (46.5) |
| Cigarettes | all[b] | 10 114 | 10 696 | (30.0) | 2161 | (42.8) | 1106 | (52.2) | 625 | (56.5) | 319 | (59.3) | 653 | (55.4) |
| | exclusive[c] | 5411 | 10 350 | (29.6) | 1472 | (39.5) | 299 | (44.4) | 100 | (49.3) | 32 | (42.7) | 355 | (52.7) |
| Snus | all[b] | 3834 | 13 373 | (32.5) | 1091 | (48.1) | 232 | (56.7) | 123 | (58.6) | 165 | (56.9) | 347 | (56.5) |
| | exclusive[c] | 1372 | 11 731 | (30.7) | 445 | (42.1) | 33 | (55.0) | 11 | (47.8) | 13 | (50.0) | 96 | (49.7) |
| **Upper secondary** | | | | | | | | | | | | | | |

| | | | | | | | | |
|---|---|---|---|---|---|---|---|---|
| Boys | | | | | | | | |
| Nicotine pouches | all[b] | 12 108 | 1479 (7.6) | 266 (6.9) | 77 (8.1) | 111 (8.0) | 448 (9.4) | 85 (8.9) |
| | exclusive[c] | 8349 | 1444 (7.5) | 216 (6.2) | 39 (6.2) | 40 (5.7) | 195 (7.4) | 71 (8.9) |
| E-cigarettes | all[b] | 14 244 | 1292 (7.4) | 533 (7.0) | 160 (8.5) | 107 (11.1) | 161 (17.6) | 267 (10.0) |
| | exclusive[c] | 7906 | 1235 (7.2) | 406 (6.9) | 51 (8.3) | 25 (13.4) | 33 (14.9) | 111 (11.8) |
| Cigarettes | all[b] | 13 617 | 1336 (7.2) | 453 (7.1) | 254 (8.2) | 162 (11.4) | 153 (16.9) | 169 (10.6) |
| | exclusive[c] | 7255 | 1286 (7.1) | 361 (7.4) | 80 (7.3) | 35 (10.8) | 26 (13.6) | 90 (13.4) |
| Snus | all[b] | 9434 | 1668 (7.6) | 333 (7.6) | 95 (8.2) | 73 (12.5) | 147 (11.1) | 167 (8.9) |
| | exclusive[c] | 3725 | 1534 (7.4) | 187 (6.9) | 12 (5.7) | 6 (13.0) | 19 (11.2) | 41 (7.4) |
| Girls | | | | | | | | |
| Nicotine pouches | all[b] | 9638 | 7490 (27.9) | 1394 (34.0) | 380 (38.3) | 437 (39.5) | 1190 (44.7) | 291 (41.1) |
| | exclusive[c] | 6244 | 7050 (27.3) | 945 (30.5) | 148 (30.6) | 189 (35.1) | 650 (40.8) | 178 (36.3) |
| E-cigarettes | all[b] | 17 717 | 5039 (26.0) | 2799 (31.1) | 836 (37.3) | 628 (42.8) | 767 (47.7) | 1274 (38.9) |
| | exclusive[c] | 12 844 | 4975 (25.9) | 2389 (29.8) | 419 (31.9) | 275 (38.2) | 406 (44.4) | 674 (37.2) |
| Cigarettes | all[b] | 14 467 | 5928 (26.2) | 2378 (32.5) | 1249 (39.5) | 602 (46.3) | 356 (49.1) | 814 (44.0) |
| | exclusive[c] | 9018 | 5777 (26.0) | 1791 (30.9) | 530 (35.1) | 198 (40.7) | 141 (47.6) | 363 (41.8) |
| Snus | all[b] | 5299 | 9059 (29.1) | 1237 (37.4) | 201 (44.7) | 81 (47.4) | 133 (44.9) | 449 (43.6) |
| | exclusive[c] | 2037 | 7740 (27.7) | 493 (29.7) | 20 (32.3) | 3 (23.1) | 16 (29.6) | 83 (36.2) |

[a] Number of cases ever used the given tobacco and nicotine product (including those tried once or twice, using currently, and quitters).

[b] Including all cases using the product.

[c] Excluding those using weekly or daily any other tobacco and nicotine products.
